# Ambient Air Pollution and Subclinical Cardiovascular Disease Measured by Magnetic Resonance Imaging in the Canadian Alliance for Healthy Hearts and Minds Study

**DOI:** 10.1101/2022.08.29.22279358

**Authors:** Sandi M Azab, Karleen M Schulze, Jeffrey R Brook, Dany Doiron, Eric E Smith, Alan R Moody, Dipika Desai, Michael Brauer, Matthias G Friedrich, Shrikant I Bangdiwala, Dena Zeraatkar, Douglas Lee, Sonia S Anand, Russell J de Souza

## Abstract

**Background:** Long-term exposure to air pollution, even at levels below regulatory standards, has been associated with higher risk of cardiovascular-related mortality. Less is known about the association of air pollution and initial development of CVD in low-exposure settings in generally healthy human populations.

**Objective:** In the Canadian Alliance for Healthy Hearts and Minds Cohort Study (CAHHM), we aimed to investigate the association between low-level exposure to key air pollutants and subclinical carotid atherosclerosis in adults without known clinical CVD. To date, the association of ambient air pollution and atherosclerosis measured by magnetic resonance imaging (MRI) has not been studied.

**Methods:** We studied 6,645 Canadian adults recruited between 2014-2018 from the provinces of British Columbia, Alberta, Ontario, Quebec, and Nova Scotia, for whom average long-term exposures to nitrogen dioxide (NO_2_), ozone (O_3_), and fine particulate matter (PM_2.5_) were estimated for five years prior to the start of CAHHM recruitment, and who underwent MRI to assess carotid vessel wall volume (CWV). Linear mixed models were used to quantify associations between each air pollutant and CWV adjusting for individual-level and community-level risk factors for CVD. Secondary analyses included region-specific stratification and modeling the effect of one pollutant on CWV within low, medium, and high levels of a second pollutant to test for interactions.

**Results:** Higher PM_2.5_ was nominally associated with lower CWV (quintile 5: 893.3 mm^3^, quintile 1: 908.8 mm^3^; *p-trend* =0.05), but this was not robust in region-stratified analysis. Higher NO_2_ was associated with lower CWV (quintile 5: 889.5 mm^3^, quintile 1: 918.6 mm^3^; *p-trend* <.0001). Higher O_3_ was associated with higher CWV (quintile 5: 925.4 mm^3^, quintile 1: 899.7 mm^3^; *p-trend* =0.02). NO_2_ emerged as a consistent effect modifier of both PM_2.5_ and O_3._

**Conclusion:** In a cohort of generally healthy adults living in Canada, a country with relatively low levels of air pollution, exposure to NO_2_ was negatively associated, and O_3_ was positively associated with CWV as a measure of subclinical atherosclerosis by MRI, while associations to PM_2.5_ were inconsistent. The reasons for these associations warrant further study.

## Introduction

Cardiovascular disease (CVD) is a leading cause of mortality in Canada and worldwide, and traditional risk factors include smoking, abdominal obesity, diabetes, hypertension, and dyslipidemia.^1,2,3^ An extensive body of literature describes the adverse effects of air pollution on cardiovascular health.^4,5,6,7,8^ It is therefore of paramount importance to ascertain the role of air pollution on early CVD indicators, such as preclinical cardiac and vascular dysfunction.

Major pollutants include particulate matter (PM) and gaseous air pollutants such as nitrogen oxides (NO_x_) and ozone (O_3_).^9^ Fine particulate matter (PM_2.5_) is fine inhalable particles ≤2.5 μm in aerodynamic diameter. Nitrogen dioxide (NO_2_) is mainly associated with road traffic and other forms of combustion and is a precursor to O_3_ ^10, 11^ A recent systematic review and meta-analysis including 353 studies of long-term exposure to traffic-related air pollution (TRAP) assessed the overall confidence in the evidence for its association with all-cause, circulatory, and ischemic heart disease (IHD) mortality as high, with non-fatal IHD and diabetes as moderate, and with coronary events as low.^12^

Studying subclinical markers of cardiovascular dysfunction, *e*.*g*., atherosclersis, is an important step in advancing our understanding of how air pollution impacts vascular health - in addition to its established effects on traditional cardiovascular risk factors,^13^ *e*.*g*., insulin resistance, hypertension, and obesity.^14,15^ The association of air pollution with early CVD development/initiation may be mediated through chronic effects on atherosclerosis progression, or possibly by trigerring acute cardiovascular events.^16^

Imaging of the carotid arteries is a noninvasive biomarker of subclinical atherosclerosis for early prediction of IHD risk in healthy individuals without CVD.^11^ In the Multiethnic Study of Atherosclerosis (MESA), outdoor O_3_ was associated with increased progression of carotid intima-media thickness (cIMT), while PM_2.5,_ NO_2,_ and NO_X_ were not associated with cIMT change.^5,11^ Magnetic resonance imaging (MRI) can accurately assess the presence of subclinical cerebrovascular atherosclerosis^17^ by measuring carotid vessel wall volume (CWV) i.e. the entire circumference of the wall. MRI-determined CWV, compared to ultrasound-measured cIMT, includes the adventitia, which is the source of vasa vasorum that further proliferates with arterial wall thickening.^18^ Thus, CWV may be a more sensitive measure of early plaque.^18^ To date, the association of ambient air pollution and MRI-captured CWV has not been studied. In the Canadian Alliance for Healthy Hearts and Minds Cohort Study (CAHHM)^19^ of generally healthy adults, we sought to investigate the associations between exposure to PM_2.5_, NO_2,_ and O_3_ in a low-level setting, and MRI-measured carotid atherosclerosis as a major CVD pathway.

## Methods

### Study design and participants

The design and methods of the CAHHM prospective cohort study have been previously described.^19^ Briefly, participants were recruited in 2014-2018 from the provinces of British Columbia, Alberta, Ontario, Quebec, and Nova Scotia in mostly urban locations.^19^ The cohort includes 8,258 adults from across Canada, of whom > 80% were participants in ongoing prospective cohort studies and phenotyped for CVD risk factors. MRI scans of the brain, heart, carotid artery, and abdomen were performed at enrollment, and 7973 participants completed a standard carotid MRI scan. Adults with known history of CVD, incomplete data on the non-lab based cardiovascular risk score, or incomplete air pollutants values were excluded for the presented analyses, leaving a final sample size of 6645 participants (**Figure S1**).

The study was approved by the Hamilton Integrated Research Ethics Board (HiREB # 13-255), and all participants provided written informed consent.

### Assessment of air pollution exposure

The three major air pollutants of interest in this study are PM_2.5_, NO_2_, and O_3_. The development of these exposure datasets has been documented elsewhere^20,21,22,23^ and they have been used in multiple Canadian epidemiological studies^24^, including the recent Mortality-Air Pollution Associations in Low Exposure Environments (MAPLE) study.^25^ Briefly, annual average exposures for the five years prior to the start of CAHHM recruitment (2008-2012), data distributed by the Canadian Urban Environmental Health Research Consortium (CANUE)^26^ (www.canue.ca), were linked to CAHHM using the six-character residential postal code of participants at the time of recruitment. The average exposure over the five years prior to recruitment is considered to be representative of long-term spatial air pollution exposure gradients and relevant for investigating subclinical CVD, which manifests over a long period of time.^27^

Key emission sources for PM_2.5_ are industry, wildfire smoke, space heating, residential wood heating, cooking, agriculture and vehicle traffic. A significant fraction of PM_2.5_ is a result of atmospheric chemistry, forming from a range of gaseous precursors such as sulphur dioxide (SO_2_), NO_2_, ammonia (NH_4_), and volatile and semivolatile organic compounds. Yearly averages of PM_2.5_ concentrations prior to baseline assessment were estimated across a 1×1 kilometer grid covering North America using NASA MODIS, MISR, and SeaWIFS satellite instruments, with aerosol vertical profiles and scattering properties simulated by the GEOS-Chem chemical transport model.^20^ To adjust for any residual bias in the satellite-derived PM_2.5_ estimates, a geographically weighted regression (GWR) incorporating ground-based observations was then applied.^20^ Good agreement was found with cross-validated surface observations across North-America (R^2^ = 0.70).

NO_2_ is considered an indicator of TRAP, which is a complex mixture of gases and particles, including ultrafine particles (diameter of 0.1 µm or less). Annual average NO_2_ concentrations in parts per billion (ppb) were estimated for each postal code location using a national land use regression (LUR) model for the year 2006^21^ at 100 m resolution and adjusted for prior and subsequent years using long-term air quality monitoring station data. The LUR NO_2_ model included road length, 2005-2011 satellite NO_2_ estimates, area of industrial land use within 2 km, and summer rainfall as predictors of regional NO_2_ variation.^21^ Deterministic gradients were used to model local scale variation related to roads (i.e., traffic).^21^ The final NO_2_ model showed good performance, explaining 73% of the variation in measurements from national air pollution surveillance (NAPS) monitoring data with a root mean square error (RMSE) of 2.9 ppb.

O_3_ is a photochemically produced oxidant gas that results from the reaction between sunlight and nitrogen oxides (NOx) and volatile organic compounds (VOCs) emitted from various natural sources and human activities such as fossil fuel combustion and wood combustion. Annual mean concentrations of O_3_ exposure at 10-15 km resolution were estimated using the GEM-MACH (Global Environmental Multi-scale - Modelling Air Quality and Chemistry) air quality forecast model combined with observations from monitoring networks.^22,23,28^

### Subclinical MRI outcomes

Details of the CAHHM MRI protocol have been previously published.^19^ The protocol used validated standard techniques to collect information on morphology, function and tissue characteristics. Briefly, participants underwent a short non-contrast enhanced scan using a 1.5 or 3.0 Tesla magnet. Each of the centres underwent a validated test scan for quality assurance. Carotid artery vessel wall volume (mm^3^) (left, right, and combined) within a 32-mm vessel length centred on each carotid bifurcation (to include distal common and proximal internal carotid arteries) was measured by subtracting lumen volume from total vessel volume. We used the maximum of either the left or right CWV as the measure of atherosclerosis in this study.

### The INTERHEART risk score

The non-laboratory-based INTERHEART risk score is a validated tool developed to estimate a person’s MI risk based on a compilation of risk factors.^29^ These include age, sex, smoking, second-hand smoke exposure, diabetes, high blood pressure, and family history of MI, waist-to-hip ratio (WHR); home or work social stress, depression, simple dietary questions, and physical activity.^2^ The score ranges from 0 to 48 and is categorized into low- (a score of 9 or lower), moderate- (a score of 10 to 15), or high- (a score of 16 or higher) risk categories and is significantly associated with diagnosed CVD, and also the presence of subclinical cerebrovascular disease without known clinical CVD.^17,29^

### Definitions

Neighbourhood walkability: Participants residential postal codes were linked to the Canadian Active Living Environments Index (Can-ALE) categorical variable that characterizes the favourability/friendliness of active living (i.e. walkability) potential of neighbourhoods in census metropolitan areas (CMA) on a scale from 1 (very low) to 5 (very high) based on intersection density, dwelling density, and points of interest measures available for the year 2016. An environment with a very high walkability tends to be densely populated and has very connected street patterns and a variety of walking destinations. More information on Can-ALE is available at: http://canue.ca/wp-content/uploads/2018/03/CanALE_UserGuide.pdf.

Rural/urban residence: Status was defined as “urban” if 21% or more postal codes within the participant’s forward sortation area (FSA) fell within a CMA or a census agglomeration (CA), and “rural” if ≤ 20% of postal codes fell within a CMA/CA.

### Statistical analysis

The distribution of continuous variables are presented as means with standard deviation, and categorical variables are presented as counts and percentages. The association of air pollutants with CWV was assessed using two approaches. First, air pollutant values were placed into quintiles, and we assessed the difference in CWV between quintiles of exposure, and the trend across quintiles. Second, we assessed the association between the continuous measure of air pollutant exposure with CWV, expressed as a 5 μg/m^3^ increment for PM_2.5_, a 5 ppb increment for NO_2_, and a 3 ppb increment for O_3_, as previously reported for relevant air pollutant concentrations in developed countries.^11,12^

The associations were explored using 5 linear mixed models (GLMM), each with random intercepts representing the effect of recruitment centre. This random intercept was also a good proxy for region of the country because most participants resided in major cities. The following fixed covariates were included in each model: (1) none (“unadjusted model”); (2) participant’s age, sex, and ethnicity (“basic model”); (3) further adjusted for contributing individual-level factors *i*.*e*., the INTERHEART risk score and education (“lifestyle model”); **(4) further adjusted for community-level factors i.e. walkability (our a priori “primary model”)** and (5) model further adjusted for co-pollutants within two-pollutant models at a time (“co-pollutant model”). In sensitivity analysis, model 4 additionally adjusted for material factor score and social factor score that together capture neighborhood socioeconomic status. A complete case analysis was employed because missing data on covariates was low (1.6% for education and 0.08% for Can-ALE index). CWV is presented as adjusted least square means (95% CI) with the p-value for trend across quintiles of exposures calculated using linear contrasts. A 2-sided p <0.05 was considered nominally significant with no adjustment for multiple testing.

In secondary analyses, a region-specific stratified analysis was conducted to consider inter-region variability. Here, we included only participants with urban residence and studied the pollutant-CWV associations for urban Canada overall and for each of the 5 regions/provinces separately. Estimates of the change in CWV (95% CI) for the specified unit change in continuous pollutant measures were presented with the same fixed covariates of models 1-5. To investigate interactions between the pollutants, we modeled the effect of one pollutant on CWV within low, medium, and high levels of a second pollutant with the same fixed covariates of models 1-4 and tested the statistical significance of the interaction term of the two pollutants. All analyses were completed using SAS software, version 9.4 (SAS Institute Inc).

## Results

### Participant characteristics

Of the 6645 participants enrolled in this study, 3253 (48.9%) were from Ontario, 1575 (23.7%) from Quebec, 744 (11.2%) from British Columbia, 671 (10.1%) from Nova Scotia, and 402 (6.0%) from Alberta. For all regions, over 92% of the cohort’s postal codes were in urban areas. The mean age of participants at enrolment was 57.6 years (SD = 8.8) and 54.8% of participants were women. The mean CWV of participants was 900.1 mm^3^ (165.1) at the time of the MRI scan. Demographic, anthropometric, and lifestyle characteristics of the study participants are found in **Tables 1 and 2**. The mean (SD) 5-year pollutant concentrations immediately preceding enrolment for PM_2.5_ was 6.9 μg/m³ (2.0), ranging from the lowest [3.2 μg/m³ (0.5)] in parts of the Calgary, Alberta, region to the highest [8.6 μg/m³ (1.5)] in London, Ontario; for NO_2_ was 12.9 ppb (5.9), ranging from lowest [4.1 ppb (1.2)] in Halifax, Nova Scotia to highest [17.0 ppb (3.9)] in Toronto, Ontario; and for O_3_ was 24.6 ppb (4.0), ranging from lowest [16.9 ppb (3.0)] in Vancouver, British Columbia to highest [30.4 ppb (1.2)] in London, Ontario, as presented in **Table 3**.

**Table 1.** Anthropometric characteristics of the study population by region of Canada.

|  |  |  | Region of Canada |  |  |  |  |
| --- | --- | --- | --- | --- | --- | --- | --- |
|  | N | Overall | BC | AB | ON | PQ | NS |
| Number of participants | 6645 | 6645 | 744 | 402 | 3253 | 1575 | 671 |
| Women, n (%) | 6645 | 3718 (56.0) | 408 (54.8) | 197 (49.0) | 1948 (59.9) | 811 (51.5) | 354 (52.8) |
| Age, y | 6645 | 57.6 (8.8) | 56.8 (8.8) | 53.3 (8.8) | 57.5 (9.0) | 58.6 (7.8) | 59.0 (9.3) |
| Weight, kg | 6645 | 76.3 (16.5) | 72.5 (15.9) | 80.7 (17.5) | 75.7 (16.5) | 77.4 (16.4) | 78.1 (15.4) |
| Height, cm | 6645 | 168.5 (9.4) | 167.5 (9.6) | 173.2 (10.1) | 168.3 (9.1) | 167.6 (9.3) | 169.4 (9.2) |
| **Body Mass Index, mean (SD), kg/m<sup>2</sup>** | 6645 | 26.8 (4.9) | 25.7 (4.4) | 26.8 (4.9) | 26.6 (4.9) | 27.5 (5.0) | 27.2 (4.8) |
| <25 (Normal), n (%) | 6645 | 2657 (40.0) | 352 (47.3) | 163 (40.5) | 1366 (42.0) | 535 (34.0) | 241 (35.9) |
| 25-29 (Overweight), n (%) | 6645 | 2536 (38.2) | 285 (38.3) | 150 (37.3) | 1193 (36.7) | 647 (41.1) | 261 (38.9) |
| 30+ (Obese), n (%) | 6645 | 1452 (21.9) | 107 (14.4) | 89 (22.1) | 694 (21.3) | 393 (25.0) | 169 (25.2) |
| Percent Body Fat, % | 6624 | 30.6 (9.2) | 28.9 (8.4) | 29.9 (9.1) | 30.9 (9.4) | 31.3 (8.8) | 30.0 (9.6) |
| Waist, cm | 6645 | 88.6 (13.7) | 84.3 (12.8) | 91.0 (13.9) | 87.5 (13.6) | 90.3 (14.0) | 93.0 (12.5) |
| Hip, cm | 6645 | 101.3 (10.7) | 99.2 (9.2) | 103.6 (10.3) | 100.4 (11.5) | 102.3 (9.7) | 104.7 (9.2) |
| Waist to hip ratio | 6645 | 0.87 (0.08) | 0.85 (0.09) | 0.88 (0.08) | 0.87 (0.08) | 0.88 (0.09) | 0.89 (0.08) |
| Waist circumference obese, n (%) | 6645 | 1924 (29.0) | 118 (15.9) | 130 (32.3) | 917 (28.2) | 492 (31.2) | 267 (39.8) |
| **Blood Pressure** |  |  |  |  |  |  |  |
| Systolic, mmHg | 6645 | 129 (17) | 126 (17) | 127 (14) | 128 (17) | 131 (16) | 133 (16) |
| Diastolic, mmHg | 6645 | 80 (10) | 80 (10) | 85 (9) | 78 (10) | 80 (9) | 80 (10) |
| Heart rate, beats/minute | 6644 | 70.4 (11.0) | 69.4 (11.0) | 70.5 (11.0) | 71.1 (11.1) | 69.7 (10.7) | 69.6 (10.6) |
| **MRI-measured Outcomes** |  |  |  |  |  |  |  |
| Carotid wall volume, mm<sup>3</sup> | 6645 | 900.1 (165.1) | 897.9 (170.5) | 889.9 (170.5) | 906.0 (163.2) | 893.2 (155.8) | 896.2 (184.3) |

Presented data are means (SD) unless otherwise indicated.

**Table 2.** Demographics & Lifestyle characteristics of the study population by region of Canada.

|  |  |  | Region of Canada |  |  |  |  |
| --- | --- | --- | --- | --- | --- | --- | --- |
|  | N | Overall | BC | AB | ON | PQ | NS |
| Number of participants | 6645 | 6645 | 744 | 402 | 3253 | 1575 | 671 |
| Women | 6645 | 3718 (56.0) | 408 (54.8) | 197 (49.0) | 1948 (59.9) | 811 (51.5) | 354 (52.8) |
| Age, mean (SD), y | 6645 | 57.6 (8.8) | 56.8 (8.8) | 53.3 (8.8) | 57.5 (9.0) | 58.6 (7.8) | 59.0 (9.3) |
| **Self-reported ethnicity** |  |  |  |  |  |  |  |
| East & South East Asian | 6645 | 894 (13.5) | 282 (37.9) | 14 (3.5) | 585 (18.0) | 8 (0.5) | 5 (0.7) |
| South Asian | 6645 | 223 (3.4) | 67 (9.0) | 3 (0.7) | 148 (4.5) | 1 (0.1) | 4 (0.6) |
| White | 6645 | 5387 (81.1) | 364 (48.9) | 381 (94.8) | 2449 (75.3) | 1545 (98.1) | 648 (96.6) |
| Other<sup>a</sup> | 6645 | 141 (2.1) | 31 (4.2) | 4 (1.0) | 71 (2.2) | 21 (1.3) | 14 (2.1) |
| **Highest Education Attained** |  |  |  |  |  |  |  |
| High school or less | 6541 | 842 (12.9) | 100 (13.5) | 44 (10.9) | 344 (10.6) | 290 (18.4) | 64 (10.9) |
| College or Trade | 6541 | 2107 (32.2) | 255 (34.5) | 117 (29.1) | 891 (27.5) | 671 (42.6) | 173 (29.4) |
| University Degree | 6541 | 3592 (54.9) | 385 (52.0) | 241 (60.0) | 2001 (61.8) | 613 (38.9) | 352 (59.8) |
| **Smoke status** |  |  |  |  |  |  |  |
| Current | 6645 | 352 (5.3) | 29 (3.9) | 18 (4.5) | 156 (4.8) | 119 (7.6) | 30 (4.5) |
| Former | 6645 | 2241 (33.7) | 178 (23.9) | 132 (32.8) | 971 (29.8) | 732 (46.5) | 228 (34.0) |
| Never | 6645 | 4052 (61.0) | 537 (72.2) | 252 (62.7) | 2126 (65.4) | 724 (46.0) | 413 (61.5) |
| **Living with partner/married** | 6537 | 4938 (75.5) | 567 (76.6) | 314 (78.1) | 2431 (75.2) | 1141 (72.4) | 485 (82.5) |
| **Employment** |  |  |  |  |  |  |  |
| Full or part time | 6534 | 4624 (70.8) | 554 (74.7) | 324 (80.6) | 2208 (68.2) | 1124 (71.5) | 414 (71.4) |
| Retired | 6534 | 1436 (22.0) | 143 (19.3) | 41 (10.2) | 727 (22.5) | 385 (24.5) | 140 (24.1) |
| No paid work | 6534 | 474 (7.3) | 45 (6.1) | 37 (9.2) | 302 (9.3) | 64 (4.1) | 26 (4.5) |
| **Individual Social disadvantage score** |  |  |  |  |  |  |  |
| Social disadvantage Index, mean (SD) | 6121 | 1.2 (1.3) | 1.2 (1.3) | 0.8 (1.1) | 1.2 (1.3) | 1.4 (1.3) | 1.1 (1.3) |
| Low disadvantage | 6121 | 3677 (60.1) | 435 (63.0) | 283 (73.7) | 1758 (59.1) | 858 (56.9) | 343 (61.0) |
| Moderate disadvantage | 6121 | 2072 (33.9) | 209 (30.3) | 92 (24.0) | 1031 (34.6) | 542 (35.9) | 198 (35.2) |
| High disadvantage | 6121 | 372 (6.1) | 46 (6.7) | 9 (2.3) | 187 (6.3) | 109 (7.2) | 21 (3.7) |
| **INTERHEART risk score mean(SD)** | 6645 | 10.0 (5.7) | 9.2 (5.5) | 9.6 (5.7) | 10.1 (5.7) | 10.3 (5.8) | 10.2 (5.6) |
| Low | 6645 | 3392 (51.0) | 418 (56.2) | 217 (54.0) | 1645 (50.6) | 785 (49.8) | 327 (48.7) |
| Moderate | 6645 | 2123 (31.9) | 223 (30.0) | 121 (30.1) | 1069 (32.9) | 491 (31.2) | 219 (32.6) |
| High | 6645 | 1130 (17.0) | 103 (13.8) | 64 (15.9) | 539 (16.6) | 299 (19.0) | 125 (18.6) |

Presented data are n (%) unless otherwise specified. <sup>a</sup> Includes Blacks, Indigenous, Mixed and unknown ethnicity

**Table 3.**
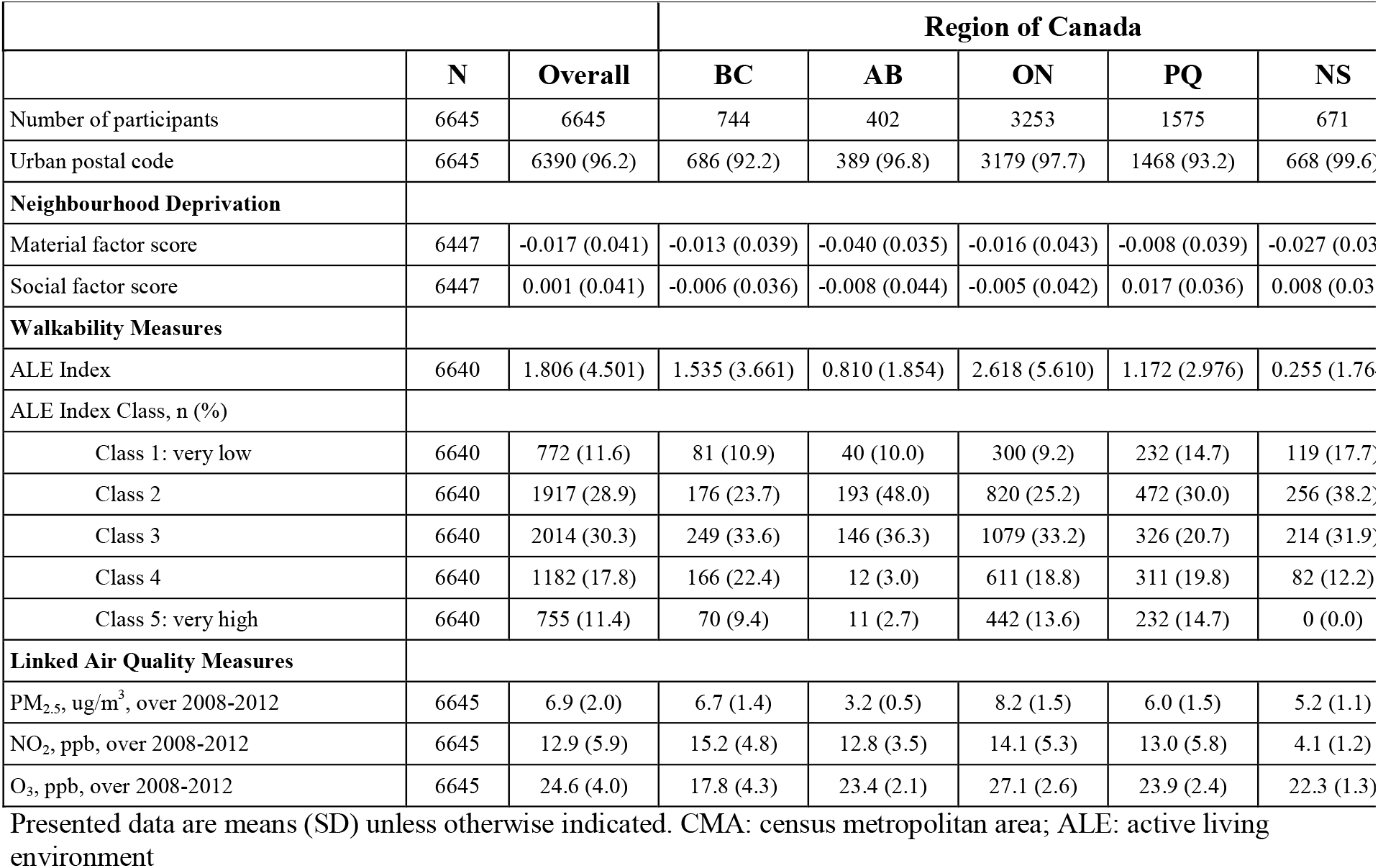
Environmental characteristics of the study population by region of Canada.

### Long-term pollutant exposure and MRI-measured CWV - Primary analysis

Associations of 5-year pollutant exposures with subclinical atherosclerosis as measured by CWV are presented in **Figure 1** and **Table S1**. An inverse association of PM_2.5_ quintile with CWV was observed (Model 4; quintile 5: 893.3 mm^3^, quintile 1: 908.8 mm^3^; *p-trend* = 0.05) which was not robust to further adjustment i.e., co-pollutant model. CWV was lower across quintiles of NO_2_ (Model 4; quintile 5: 889.5 mm^3^, quintile 1: 918.6 mm^3^; *p-trend* < 0.0001) in all models including those with co-pollutant adjustment, but was higher across quintiles of O_3_ (quintile 5: 925.4mm^3^, quintile 1: 899.7; *p-trend* = 0.02), which was blunted after NO_2_ but not PM_2.5_ adjustment. These results remained consistent in the sensitivity analysis (Table S6). Of note, Pearson correlation between PM_2.5_ and NO_2_ was [r=0.39; p<.0001], between PM_2.5_ and O_3_ was [r=0.16; p<.0001], and between NO_2_ and O_3_ was [r=-0.23; p<.0001].

**Figure 1:**
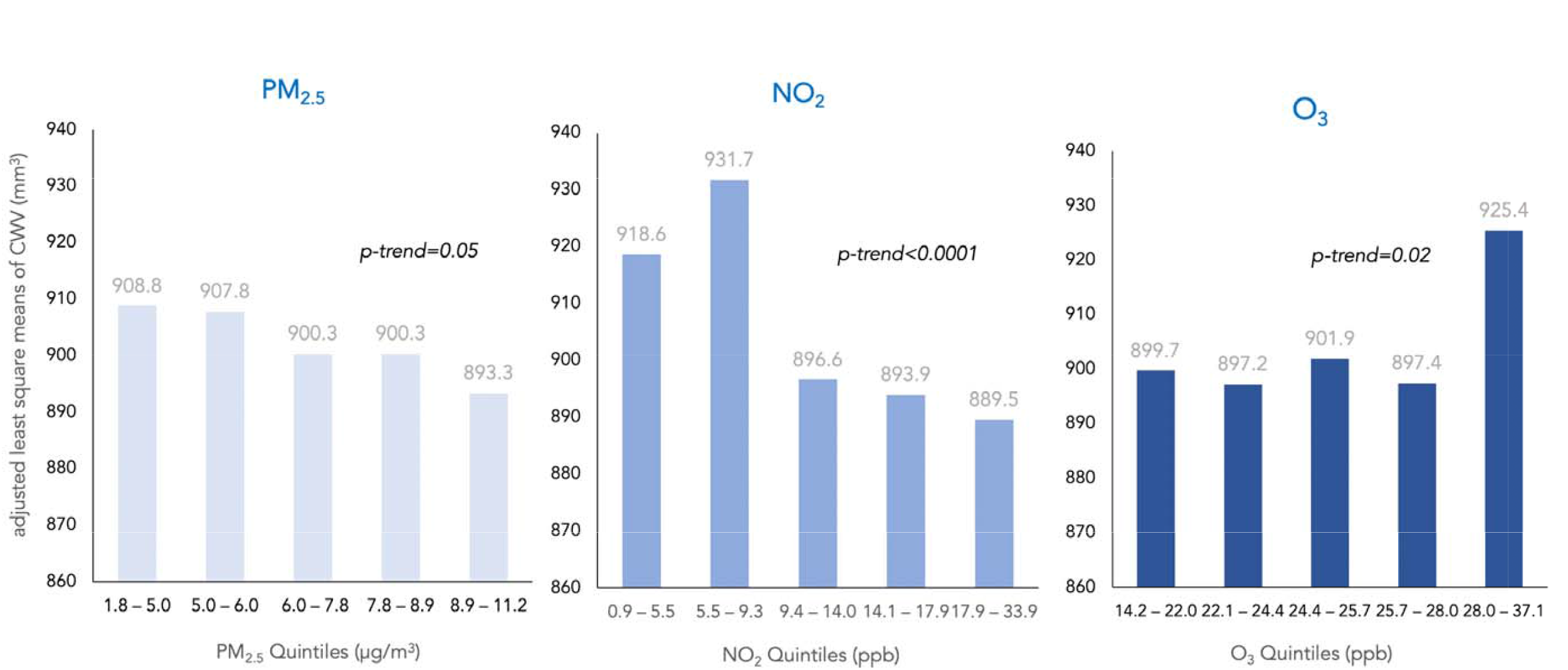
Associations of 5-year pollutant exposures with subclinical atherosclerosis as measured by carotid wall volume (CWV)

### Region-specific association of air pollutants and CWV

In a secondary region-stratified analysis across urban Canada (n=6282), overall trends observed in the primary analysis for the three pollutants were replicated using continuous measures for the air pollutants (**Figure 2** and **Table S2**). The associations between PM_2.5_ and CWV was not statistically significant in urban Canada and varied across provinces. Within Ontario, after adjustment for neighbourhood walkability in the primary model (Model 4, table 5) the negative association between PM_2.5_ and CWV was no longer seen. Only when further adjusting for NO_2_ in the co-pollutant model, a 5 μg/m^3^ higher PM_2.5_ concentration was associated with 24.98 mm^3^ higher CWV (95% CI 3.43-46.52; p=0.023). A 5 ppb higher NO_2_ concentration was associated with 21.2 mm^3^ lower CWV (95% CI −27.7--14.7; p<0.0001) within Ontario (n=3161), but with a 100.6 mm^3^ higher CWV (95% CI 24.99-176.2; p=0.009) in Nova Scotia (n=586). A 3 ppb increase in O_3_ was associated with 9.66 mm^3^ (95% CI 1.69-17.63; p=0.018) and 11.01 mm^3^ (95% CI 0.56-21.47; p=0.039) higher CWV in Ontario and Quebec (n=1466), respectively; the two regions with largest sample size.

**Figure 2:**
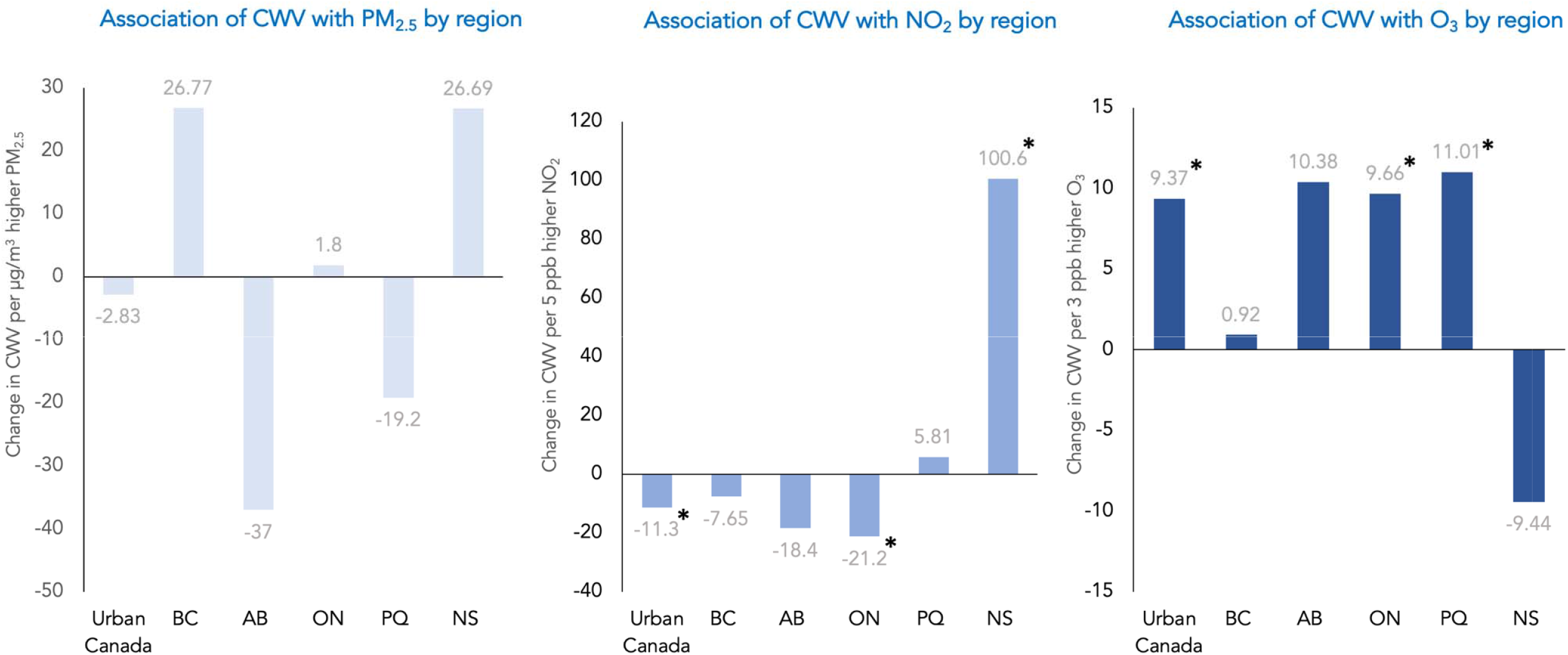
Region-specific association of air pollutants and carotid wall volume (CWV) across urban Canada. *p<0.05

### Effect of co-pollutant interactions on CWV

Across the primary models summarized in **Figure 3** for testing the effect of one pollutant on CWV within low, medium, and high levels of a second pollutant, the effect of PM_2.5_ on CWV depended on the level of exposure to NO_2_ (*p=0*.*014* for interaction). There was no evidence of interaction between PM_2.5_ and O_3_. Furthermore, the effect of NO_2_ on CWV differed according to both the exposure levels of PM_2.5_ (*p=0*.*024* for interaction) and O_3_ (*p<0*.*0001* for interaction). The effect of O_3_ on CWV depended on NO_2_ levels (*p=0*.*0006* for interaction). Thus, NO_2_ emerged as a consistent effect modifier of both PM_2.5_ and O_3_ (**Tables S3–S5**).

**Figure 3:**
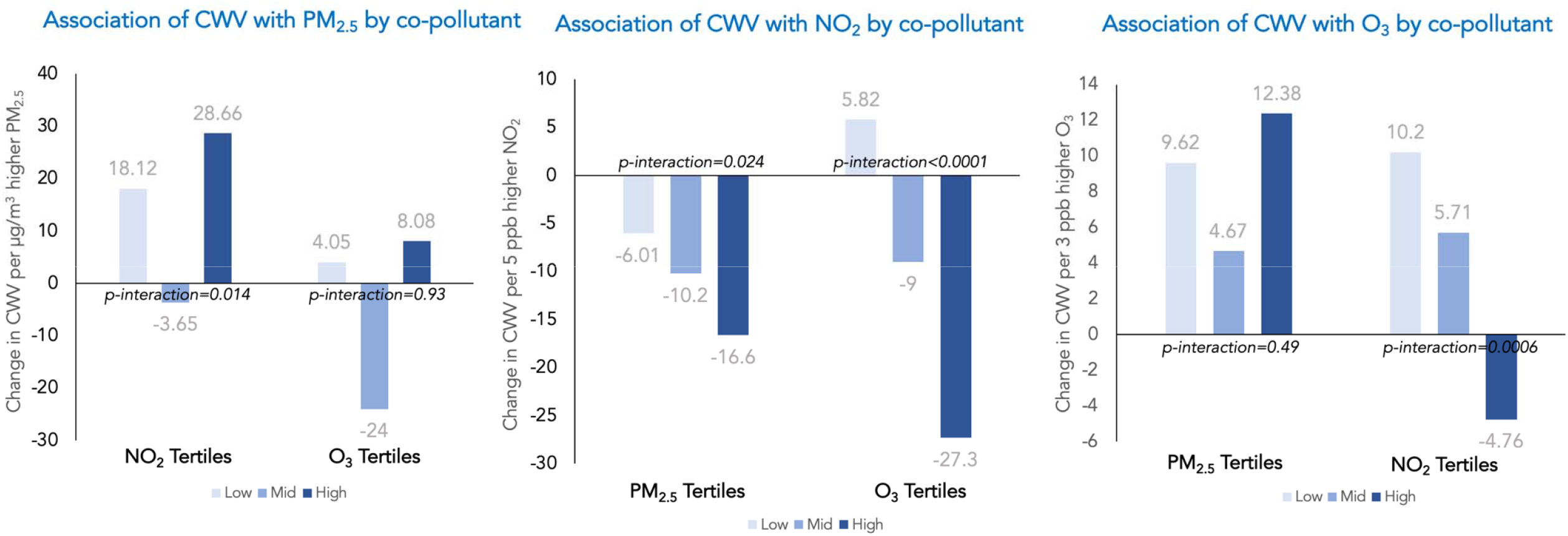
Effect of copollutant interactions on carotid wall volume (CWV)

## Discussion

In this prospective cohort study among 6,645 Canadian adults and using spatially resolved pollutant concentrations, long-term exposure to air pollution was consistently associated with decreased MRI-measured CWV for NO_2,_ increased MRI-measured CWV for O_3_, and for a trend towards decreased MRI-mesured CWV for PM_2.5_. While findings for NO_2_ and O_3_ were robust to some extent in secondary analyses, results for PM_2.5_ were not. Effects beyond adjusting for the INTERHEART risk score suggest that air pollution is possibly also acting through other pathways than the traditional cardiovascular risk factors captured in the score *e*.*g*., diabetes and hypertension. NO_2_ consistently modified the effects of PM_2.5_ and O_3_ on CWV.

There is limited evidence that direct biological measures of early CVD are influenced by exposure to air pollution at low levels. However, the MAPLE study reports associations between nonaccidental mortality and cardiovascular-related mortality and long-term exposure to ambient PM_2.5_ levels, including at concentrations below national air quality standards.^25^ The evidence is moderate to strong for overall TRAP and CVD mortality, but is moderate at best for CVD morbidity *i*.*e*., the effect of air pollution on traditional cardiovascular risk factors.^12,30^ Our results show an inconsistent association between PM_2.5_ and a state-of-the-art surrogate marker for subclinical atherosclerosis across Canada, consistent with previous findings of the Multicultural Community Health Assessment Trial (M-CHAT) study between TRAP and progression of carotid artery atherosclerosis in Vancouver, BC.^16^ Though the negative association between PM_2.5_ and CWV is of borderline significance, it is small in magnitude compared with the associations seen with O_3_ and NO_2_. This suggests that PM_2.5_ may be affecting CV risk through pathways other than atherosclerosis formation or rather by affecting plaque stability. Therefore, if there is an increased risk with exposure to PM_2.5_, it is operating through other intermediary pathways such as release of proinflammatory mediators, autonomic nervous system perturbations, and translocation of particle constituents into the blood.^31^ Interestingly, neighbourhood walkability was shown to modify the effects of PM_2.5_ in the main analysis, in Ontario in the region-specific analysis, and within the high O_3_ category in the co-pollutant interaction analysis; a variable that has been scarcely captured in previous literature.

Our findings on the detrimental effect of chronic ambient O_3_ exposure on subclinical atherosclerosis are robust across regions and co-pollutant analysis. This is congruent with what the MESA study had previously reported on progression of IMT of the common carotid artery and new carotid plaque formation with outdoor O_3_ exposure in six U.S. city regions.^11^ One underlying biochemical pathway might be through the formation of reactive oxygen species (due to the strong oxidizing action of O_3_, which may have direct effects or indirect effects changing the oxidizing potential of ambient PM_2.5_) that further give rise to increased oxidative stress and persistent systemic inflammation.^11^

The weak yet significant inverse association of NO_2_, largely an indicator of TRAP and/or combustion pollution in general, with CWV in our study is unexpected and needs careful evaluation. There is insufficient collective evidence in the literature linking NO_2_ or TRAP with preclinical atherosclerosis in healthy adults specifically. A recent assessment and meta-analysis concluded that the evidence of an association between TRAP and cardiovascular morbidity is low to moderate.^12^ In fact, MESA-Air and the European Study of Cohorts for Air Pollution Effects (ESCAPE) studies did not find a relationship between NO_2_ and IMT.^5,32^ However, in a study of a high cardiovascular risk population (2227 patients mean age of 62.9 years) in London, Ontario, NO_2_ exposure was associated with cumulative plaque burden as captured by carotid total plaque area (TPA) using two-dimensional ultrasound.^33^

Importantly, our secondary analysis across Canada’s urban regions offers insight into possible regional variation in the effects of air pollution on CV health. Only in Nova Scotia (predominantly in Halifax) did we observe a positive association of NO_2_ with CWV; and Halifax was the city with the lowest levels of NO_2_ of the five Canadian cities. Of note, the ratio of O_3_/NO_2_ in Halifax is 5.3 which is much higher than the values of 1.8-1.9 observed in the other four cities/regions and in the cohort overall. Thus, our study emphasizes the need for further investigation of different exposures in combination.^8^ It also emphasizes the value of adjusting for novel neighbourhood characteristics such as active living environment, to examine effect modification and help further investigate region variability. Neighbourhood factors that might modify the association between air pollution and CWV include poverty/affluence, overcrowding, living in apartment buildings, commuting, and proximity to roads.

### Strengths and Limitations

Our study has obtained unique health measurements of subclinical cardiovascular markers using MRI on nearly 6600 Canadians along with individual-level information on environmental factors and lifestyle known to influence cardiometabolic outcomes. While IMT has been criticized as an accurate marker of atherosclerosis with sensitivity limitations of the ultrasound methodology,^33^ our study uses MRI-characterized CWV to assess atherosclerosis. Moreover, compared to MESA which reported on cIMT progression in 3392 participants with low-exposure levels, our study is well-powered. Finally, the cohort’s diverse geographic coverage across Canada offers an exposure gradient in ambient PM_2.5_, NO_2_, and O_3_ that parallels what has previously been explored by the national Canadian Census Health and Environment Cohort (CanCHEC).^25^ Several limitations are important to mention. First, individual-level exposures were estimated based on residence address. While this is common in epidemiological air pollution studies, exposure inaccuracy is inevitable due to the fact that participants’ time spent away from residence and residential history were not taken into account in estimating long-term exposure to air pollutants. Second, the 5-year pollutant exposure period was fixed for all participants (2008-2012) regardless of when an individual’s enrolment occurred in the 2014-2018 window. This exposure window was not consistently 5-years prior to enrolment for all study participants. However, studies have demonstrated temporal stability in the spatial patterns of air pollutants over 10 years.^34^ Because of the small number of events in our cohort (n=155), we were not powered to look at intraplaque hemorrhage. Lastly, our results are based on cross-sectional data therefore causality remains uncertain.

## Conclusion

Our data indicate that in healthy adults living in clean or only mildly polluted environments, exposure to NO_2_ is negatively associated, and O_3_ is positively associated with CWV as a measure of subclinical atherosclerosis. While we also observed associations to PM_2.5_, these were not consistent across regions. Especially the role of NO_2_ in atherosclerosis is complex and requires further investigation, as do combinations of exposure to O_3_, PM_2.5_ and NO_2_.

## Supporting information

Strobe Checklist

## Data Availability

All data produced in the present study are available upon reasonable request to the authors

## Acknowledgements

**Steering Committee of Canadian Alliance of Healthy Hearts and Minds:** S.S. Anand (Chair)*, M.G. Friedrich(Co-Chair), Douglas S. Lee (Co-Chair), P Awadalla (Ontario Health Study), T. Dummer (BC Generations Project), J. Vena (Alberta’s Tomorrow Project), P. Broët (CARTaGENE), J. Hicks (Atlantic PATH), J-C. Tardif (MHI Biobank), K. Teo, S. Yusuf (PURE-Central), B-M. Knoppers (Ethics, Legal and Social Issues).

**Project Office Staff at Population Health Research Institute(PHRI):** D. Desai, S. Zafar

**Statistics/BiometricsProgrammers Team at PHRI:** K. Schulze, S. Bangdiwala

**Cohort Operations Research Personnel:** K McDonald (Ontario Health Study), N. Noisel (CARTaGENE), J. Chu (BC Generations Project), J. Hicks (Atlantic PATH), H. Whelan (Alberta’s Tomorrow Project), S. Rangarajan (PURE), D. Busseuil (MHI Biobank)

**Site Investigators and Staff:** (112) J. Leipsic, S. Lear, V. de Jong; (306) M. Noseworthy, K. Teo, E. Ramezani, N. Konyer; (402) P. Poirier, A-S. Bourlaud, E. Larose, K. Bibeau;(512) J. Leipsic, S. Lear, V. de Jong; (609) E. Smith, R. Frayne, A. Charlton, R Sekhon; (703) A. Moody, V. Thayalasuthan; (704) A.Kripalani, G Leung; (706) M. Noseworthy, S. Anand, R. de Souza, N. Konyer, S. Zafar; (707) G. Paraga,L. Reid; (714) A.J. Dick, F. Ahmad; (799) D. Kelton, H. Shah; (801) F. Marcotte, H. Poiffaut; (802) M.G. Friedrich, J. Lebel; (817) E. Larose, K. Bibeau; (913) R. Miller, L. Parker, D. Thompson, J. Hicks; (1001) J-C. Tardif, H.Poiffaut; (1103) J. Tu, K. Chan, A. Moody, V. Thayalasuthan;

**MRI Working Group and Core Lab Investigators/Staff:**

**Chair:** M.G. Friedrich; **Brain Core Lab**: E.E. Smith; **Carotid Core Lab:** A. Moody, V. Thayalasuthan; **Abdomen:** E. Larose, K. Bibeau,

**Cardiac:** F. Marcotte, F. Henriques

**Contextual Working Group:** R. de Souza, S. Anand, G. Booth, J. Brook, D. Corsi, L. Gauvin, S. Lear, F. Razak, S.V. Subramanian, J. Tu.

**CAHHM Founding Advisory Group:** Jean Rouleau, Pierre Boyle, Caroline Wong, Eldon Smith

ClinicalTrials.gov, NCT02220582. Registered 20 August 2014—Retrospectively registered, https://clinicaltrials.gov/ct2/show/NCT02220582.

---

**Table S1:** Association of carotid wall volume (mm^3^) with pollutants exposure.

| Pollutant | Q1 | Q2 | Q3 | Q4 | Q5 | p-trend |
| --- | --- | --- | --- | --- | --- | --- |
| **PM<sub>2.5</sub>** |  |  |  |  |  |  |
| Quintile range | 1.8 – 5.0 ug/m<sup>3</sup> | 5.0 – 6.0 ug/m<sup>3</sup> | 6.0 – 7.8 ug/m<sup>3</sup> | 7.8 – 8.9 ug/m<sup>3</sup> | 8.9 – 11.2 ug/m<sup>3</sup> |  |
| Model 1 | 910.5 (889.4, 931.6) | 910.6 (889.4, 931.8) | 902.8 (881.9, 923.6) | 903.0 (882.0, 924.1) | 895.1 (873.7, 916.5) | 0.05 |
| Model 2 | 912.6 (888.6, 936.6) | 910.0 (886.0, 934.1) | 901.8 (878.0, 925.5) | 899.1 (875.1, 923.0) | 890.4 (866.2, 914.6) | 0.002 |
| Model 3 | 912.6 (889.0, 936.2) | 909.9 (886.1, 933.6) | 901.6 (878.1, 925.0) | 899.1 (875.4, 922.7) | 890.2 (866.3, 914.0) | 0.002 |
| Model 4 | 908.8 (885.4, 932.2) | 907.8 (884.5, 931.1) | 900.3 (877.3, 923.3) | 900.3 (877.1, 923.5) | 893.3 (869.7, 916.9) | 0.05 |
| Model 5:<br>Model4 +NO<sub>2</sub> | 905.9 (882.0, 929.7) | 905.1 (881.3, 928.8) | 897.1 (873.6, 920.6) | 904.2 (880.5, 927.9) | 897.7 (873.6, 921.8) | 0.39 |
| Model 5:<br>Model4+O<sub>3</sub> | 908.5 (888.1, 928.9) | 909.5 (889.2, 929.9) | 904.6 (884.5, 924.7) | 907.4 (886.8, 927.9) | 899.9 (879.0, 920.7) | 0.33 |
| **NO<sub>2</sub>** |  |  |  |  |  |  |
| Quintile range | 0.9 – 5.5 ppb | 5.5 – 9.3 ppb | 9.4 – 14.0 ppb | 14.1 – 17.9 ppb | 17.9 – 33.9 ppb |  |
| Model 1 | 919.2 (896.5, 941.9) | 927.2 (906.3, 948.0) | 897.4 (877.3, 917.6) | 899.8 (879.8, 919.9) | 890.7 (870.4, 911.0) | <.0001 |
| Model 2 | 918.3 (894.1, 942.6) | 929.9 (907.1, 952.7) | 895.7 (873.4, 917.9) | 894.5 (872.3, 916.8) | 890.6 (868.2, 913.0) | <.0001 |
| Model 3 | 918.5 (894.4, 942.6) | 929.6 (906.9, 952.2) | 895.4 (873.3, 917.5) | 894.6 (872.5, 916.6) | 890.9 (868.7, 913.1) | <.0001 |
| Model 4 | 918.6 (893.0, 944.2) | 931.7 (908.9, 954.6) | 896.6 (874.5, 918.7) | 893.9 (871.9, 916.0) | 889.5 (867.0, 911.9) | <.0001 |
| Model 5:<br>Model4+O<sub>3</sub> | 915.6 (891.1, 940.0) | 931.0 (909.6, 952.3) | 898.5 (877.8, 919.1) | 898.2 (877.1, 919.2) | 894.1 (872.6, 915.7) | 0.004 |
| Model 5:<br>Model4+PM<sub>2.5</sub> | 918.2 (892.1, 944.2) | 931.5 (908.5, 954.6) | 896.5 (874.3, 918.8) | 894.0 (871.7, 916.2) | 889.5 (866.9, 912.1) | <0.001 |
| **O<sub>3</sub>** |  |  |  |  |  |  |
| Quintile range | 14.2 – 22.0 ppb | 22.1 – 24.4 ppb | 24.4 – 25.7 ppb | 25.7 – 28.0 ppb | 28.0 – 37.1 ppb |  |
| Model 1 | 901.5 (882.9, 920.2) | 898.0 (879.1, 916.9) | 906.2 (887.6, 924.7) | 901.0 (882.0, 920.0) | 924.6 (905.2, 944.1) | 0.02 |
| Model 2 | 898.5 (878.8, 918.2) | 896.9 (877.0, 916.7) | 902.8 (883.1, 922.4) | 898.7 (878.7, 918.7) | 929.0 (908.7, 949.3) | 0.001 |
| Model 3 | 898.3 (878.9, 917.7) | 896.7 (877.2, 916.3) | 903.2 (883.8, 922.6) | 898.8 (879.1, 918.6) | 928.8 (908.7, 948.9) | 0.001 |
| Model 4 | 899.7 (879.7, 919.7) | 897.2 (877.2, 917.2) | 901.9 (882.0, 921.9) | 897.4 (877.0, 917.7) | 925.4 (903.9, 946.9) | 0.02 |
| Model 5:<br>Model4+NO<sub>2</sub> | 903.1 (881.2, 925.1) | 898.4 (876.5, 920.3) | 902.0 (880.2, 923.9) | 893.9 (871.6, 916.2) | 916.7 (892.9, 940.4) | 0.34 |
| Model 5:<br>Model4+PM<sub>2.5</sub> | 899.7 (879.4, 920.0) | 897.1 (876.9, 917.4) | 901.8 (881.6, 922.0) | 897.2 (876.6, 917.8) | 925.0 (903.1, 946.9) | 0.02 |

Presented data are adjusted least square means (95% CI) from a linear mixed model with centre modelled as a random intercept. Model 1, unadjusted. Model 2, adjusted for age, sex, and ethnicity. Model 3 further adjusted for the INTERHEART risk score and education. Primary model 4 further adjusted for walkability categories. Model 5 further adjusted for co-pollutants. P-trend from linear contrasts. For all models, N=6536.

**Table S2:** Association of carotid wall volume (mm^3^) with pollutants exposure across urban Canada.

|  | Urban Canada (6282) |  | BC (n=680) |  | AB (n=389) |  | ON (n=3161) |  | PQ (n=1466) |  | NS (N=586) |  |
| --- | --- | --- | --- | --- | --- | --- | --- | --- | --- | --- | --- | --- |
|  | Effect (95% CI) | p- | Effect (95% CI) | p- | Effect (95% CI) | p- | Effect (95% CI) | p- | Effect (95% CI) | p- | Effect (95% CI) | p- |
| **PM<sub>2.5</sub>** |  |  |  |  |  |  |  |  |  |  |  |  |
| Model 1 | -6.10 (-21.0,8.84) | 0.42 | 13.63 (-39.6,66.84) | 0.62 | 35.08 (-150,220) | 0.71 | -18.3 (-37.3,0.79) | 0.060 | -1.41 (-29.9,27.10) | 0.92 | -0.77 (-71.9,70.33) | 0.98 |
| Model 2 | -10.7 (-24.5,3.09) | 0.13 | 8.66 (-38.8,56.10) | 0.72 | 21.43 (-148,191) | 0.80 | -21.6 (-39.3,-3.94) | 0.016 | -8.98 (-33.2,15.20) | 0.47 | 4.15 (-60.9,69.15) | 0.90 |
| Model 3 | -11.0 (-24.8,2.79) | 0.12 | 17.10 (-30.5,64.67) | 0.48 | 12.33 (-159,184) | 0.89 | -22.0 (-39.8,-4.33) | 0.015 | -9.40 (-34.1,15.31) | 0.46 | 0.28 (-65.2,65.75) | 0.99 |
| Model 4 | -2.83 (-17.8,12.14) | 0.71 | 26.77 (-24.1,77.64) | 0.30 | -37.0 (-223,148.6) | 0.70 | 1.80 (-18.8,22.42) | 0.86 | -19.2 (-40.5,2.21) | 0.079 | 26.69 (-53.6,107.0) | 0.51 |
| Model 5:+NO<sub>2</sub> | 3.70 (-11.6,18.97) | 0.63 | 50.10 (-7.93,108.1) | 0.09 | -88.5 (-286,109.2) | 0.38 | 24.98 (3.43,46.52) | 0.023 | -18.9 (-40.3,2.45) | 0.082 | 11.27 (-69.5,92.07) | 0.78 |
| Model 5:+O<sub>3</sub> | 3.07 (-12.1,18.21) | 0.70 | 32.75 (-22.1,87.64) | 0.24 | -75.4 (-277,126.4) | 0.46 | 4.79 (-16.0,25.54) | 0.65 | -1.13 (-30.3,28.06) | 0.94 | 23.73 (-57.7,105.2) | 0.57 |
| **NO<sub>2</sub>** |  |  |  |  |  |  |  |  |  |  |  |  |
| Model 1 | -10.1 (-14.4,-5.81) | <.0001 | -11.3 (-27.7,5.17) | 0.18 | -20.6 (-46.4,5.08) | 0.12 | -20.1 (-25.7,-14.5) | <.0001 | 7.84 (0.71,14.98) | 0.031 | 42.14 (-22.2,106.5) | 0.199 |
| Model 2 | -11.1 (-15.0,-7.12) | <.0001 | -8.86 (-23.8,6.10) | 0.25 | -18.4 (-42.2,5.31) | 0.13 | -21.5 (-26.7,-16.4) | <.0001 | 6.83 (0.52,13.14) | 0.034 | 57.13 (-1.64,115.9) | 0.057 |
| Model 3 | -10.8 (-14.8,-6.85) | <.0001 | -9.20 (-24.1,5.72) | 0.23 | -16.3 (-40.2,7.68) | 0.18 | -21.4 (-26.5,-16.2) | <.0001 | 7.53 (1.07,14.00) | 0.022 | 51.46 (-8.58,111.5) | 0.093 |
| Model 4 | -11.3 (-16.3,-6.24) | <.0001 | -7.65 (-23.7,8.43) | 0.35 | -18.4 (-47.7,10.8) | 0.22 | -21.2 (-27.7,-14.7) | <.0001 | 5.81 (-4.67,16.29) | 0.28 | 100.6 (24.99,176.2) | 0.009 |
| Model 5:+O<sub>3</sub> | -8.98 (-14.5,-3.50) | 0.0013 | -9.27 (-27.8,9.23) | 0.33 | -18.5 (-54.7,17.7) | 0.32 | -22.3 (-29.6,-14.9) | <.0001 | 7.99 (-2.65,18.62) | 0.14 | 101.0 (25.38,176.7) | 0.009 |
| Model 5:+PM<sub>2.5</sub> | -11.5 (-16.6,-6.38) | <.0001 | -15.3 (-33.7,3.00) | 0.10 | -23.3 (-54.5,7.93) | 0.14 | -23.7 (-30.5,-16.8) | <.0001 | 5.43 (-5.01,15.88) | 0.31 | 99.03 (22.52,175.5) | 0.011 |
| **O<sub>3</sub>** |  |  |  |  |  |  |  |  |  |  |  |  |
| Model 1 | 8.17 (3.34,13.01) | 0.0009 | 5.57 (-6.87,18.01) | 0.38 | 8.65 (-18.1,35.41) | 0.53 | 11.75 (4.76,18.74) | 0.0010 | 1.98 (-8.30,12.26) | 0.71 | 3.05 (-31.4,37.48) | 0.86 |
| Model 2 | 10.84 (6.34,15.35) | <.0001 | 4.58 (-6.96,16.13) | 0.44 | 11.84 (-12.6,36.3) | 0.34 | 15.90 (9.47,22.32) | <.0001 | 3.38 (-5.82,12.57) | 0.47 | -2.13 (-33.8,29.59) | 0.90 |
| Model 3 | 10.66 (6.14,15.18) | <.0001 | 3.98 (-7.52,15.49) | 0.50 | 9.79 (-14.8,34.40) | 0.43 | 15.65 (9.15,22.14) | <.0001 | 3.49 (-5.81,12.79) | 0.46 | -2.83 (-34.6,28.99) | 0.86 |
| Model 4 | 9.37 (4.30,14.45) | 0.0003 | 0.92 (-11.4,13.19) | 0.88 | 10.38 (-17.9,38.7) | 0.47 | 9.66 (1.69,17.63) | 0.0176 | 11.01 (0.56,21.47) | 0.039 | -9.44 (-43.9,24.98) | 0.59 |
| Model 5:+NO<sub>2</sub> | 5.71 (0.14,11.28) | 0.0447 | -2.53 (-16.6,11.57) | 0.72 | -0.14 (-35.1,34.9) | 0.99 | -2.87 (-11.8,6.06) | 0.5284 | 12.62 (1.94,23.31) | 0.021 | -10.3 (-44.6,23.93) | 0.55 |
| Model 5:+PM<sub>2.5</sub> | 9.57 (4.41,14.74) | 0.0003 | 3.88 (-9.35,17.11) | 0.57 | 14.91 (-15.9,45.7) | 0.34 | 9.87 (1.85,17.90) | 0.0159 | 10.96 (-0.52,22.45) | 0.061 | -7.78 (-42.7,27.13) | 0.66 |

Presented data are adjusted estimates (95% CI) per 5 µg/m<sup>3</sup> increase for PM<sub>2.5</sub>, 5 ppb increase for NO<sub>2</sub>, and 3 ppb increase for O<sub>3</sub>, from a linear mixed model with centre modelled as a random intercept of region-stratified analysis. Model 1, unadjusted. Model 2, adjusted for age, sex, and ethnicity. Model 3 further adjusted for the INTERHEART risk score and education. Primary model 4 further adjusted for walkability categories. Model 5 further adjusted for co-pollutants.

**Table S3:** Association of carotid wall volume (mm^3^) with O_3_ within co-pollutant exposure tertiles.

|  | **Overall**<br>**N=6536** |  | **Low PM<sub>2.5</sub>**<br>**(1.78 - 5.64 µg/m<sup>3</sup>)**<br>**N=2040** |  | **Mid PM<sub>2.5</sub>**<br>**5.65 - 8.18**<br>**µg/m<sup>3</sup>N=1972** |  | **High PM<sub>2.5</sub>**<br>**8.18 - 11.2 µg/m<sup>3</sup>**<br>**N=2524** |  |
| --- | --- | --- | --- | --- | --- | --- | --- | --- |
|  | **Effect**<br>**(95% CI)** | **p-** | **Effect**<br>**(95% CI)** | **p-** | **Effect**<br>**(95% CI)** | **p-** | **Effect**<br>**(95% CI)** | **p-** |
| Model 1 | 7.80 (3.46,12.14) | 0.0004 | 6.63<br>(-1.40,14.67) | 0.1055 | 8.32<br>(1.00,15.64) | 0.0259 | 8.18<br>(0.24,16.13) | 0.0436 |
| Model 2 | 10.24 (6.22,14.26) | <.0001 | 10.90<br>(5.31,16.49) | 0.0001 | 5.82<br>(-1.91,13.54) | 0.1399 | 13.66<br>(6.27,21.06) | 0.0003 |
| Model 3 | 10.16 (6.13,14.20) | <.0001 | 10.77<br>(4.86,16.69) | 0.0004 | 5.80 (-<br>1.95,13.55) | 0.1424 | 12.55<br>(5.12,19.98) | 0.0009 |
| Model 4 | 9.14 (4.45,13.83) | 0.0001 | 9.62<br>(0.91,18.33) | 0.0304 | 4.67 (-<br>3.72,13.06) | 0.2747 | 12.38<br>(4.45,20.32) | 0.0022 |
|  |  |  | **Low NO<sub>2</sub>**<br>**(0.88 - 8.10 ppb)**<br>**N=1590** |  | **Mid NO<sub>2</sub>**<br>**(8.12 - 15.5 ppb)**<br>**N=2327** |  | **High NO<sub>2</sub>**<br>**(15.5 - 38.7 ppb)**<br>**N=2619** |  |
|  |  |  | **Effect**<br>**(95% CI)** | **p-** | **Effect**<br>**(95% CI)** | **p-** | **Effect**<br>**(95% CI)** | **p-** |
| Model 1 |  |  | 9.61<br>(-1.92,21.13) | 0.1022 | 0.79<br>(-6.88,8.46) | 0.8394 | -4.19<br>(-15.1,6.69) | 0.4503 |
| Model 2 |  |  | 9.90 (-<br>1.17,20.96) | 0.0796 | 4.73 (-<br>2.39,11.85) | 0.1930 | -5.04 (-<br>15.5,5.38) | 0.3430 |
| Model 3 |  |  | 10.59 (-<br>0.46,21.65) | 0.0603 | 4.87 (-<br>2.25,11.98) | 0.1802 | -5.29 (-<br>15.7,5.09) | 0.3177 |
| Model 4 |  |  | 10.20 (-<br>1.30,21.69) | 0.0821 | 5.71 (-<br>1.60,13.02) | 0.1260 | -4.76 (-<br>15.2,5.67) | 0.3711 |

Presented data are adjusted CWV estimates (95% CI) per, 3 ppb increase for O<sub>3</sub>, from a linear mixed model with centre modelled as a random intercept of regio-stratified analysis. Model 1, unadjusted. Model 2, adjusted for age, sex, and ethnicity. Model 3 further adjusted for the INTERHEART risk score and education. Primary model 4 further adjusted for walkability categories.

**Table S4:** Association of carotid wall volume (mm^3^) with PM_2.5_ within co-pollutant exposure tertiles.

|  | **Overall**<br>**N=6536** |  | **Low O<sub>3</sub>**<br>**14.2 - 22.9 ppb**<br>**N=2051** |  | **Mid O<sub>3</sub>**<br>**23.0 - 26.3 ppb**<br>**N=2422** |  | **High O<sub>3</sub>**<br>**26.3 - 39.1 ppb**<br>**N=2063** |  |
| --- | --- | --- | --- | --- | --- | --- | --- | --- |
|  | **Effect**<br>**(95% CI)** | **p-** | **Effect**<br>**(95% CI)** | **p-** | **Effect**<br>**(95% CI)** | **p-** | **Effect**<br>**(95% CI)** | **p-** |
| Model 1 | -10.6 (-25.0,3.68) | 0.1453 | 4.02 (-14.0,21.99) | 0.6610 | -6.91 (-42.1,28.26) | 0.7000 | -18.6 (-38.6,1.42) | 0.0686 |
| Model 2 | -15.7 (-28.9,-2.47) | 0.0200 | -2.16 (-27.2,22.89) | 0.8660 | -18.6 (-51.5,14.16) | 0.2651 | -18.8 (-37.3,-0.29) | 0.0466 |
| Model 3 | -15.9 (-29.1,-2.72) | 0.0181 | -0.77 (-25.9,24.33) | 0.9523 | -19.5 (-52.1,13.06) | 0.2400 | -18.9 (-37.4,-0.31) | 0.0463 |
| Model 4 | -7.94 (-22.5,6.67) | 0.2866 | 4.05 (-21.7,29.84) | 0.7578 | -24.0 (-57.7,9.59) | 0.1611 | 8.08 (-14.0,30.13) | 0.4726 |
|  |  |  | **Low NO<sub>2</sub>**<br>**(0.88 - 8.10 ppb)**<br>**N=1590** |  | **Mid NO<sub>2</sub>**<br>**(8.12 - 15.5 ppb)**<br>**N=2327** |  | **High NO<sub>2</sub>**<br>**(15.5 - 38.7 ppb)**<br>**N=2619** |  |
|  |  |  | **Effect**<br>**(95% CI)** | **p-** | **Effect**<br>**(95% CI)** | **p-** | **Effect**<br>**(95% CI)** | **p-** |
| Model 1 |  |  | 23.25 (-10.7,57.20) | 0.1793 | -0.68 (-22.4,20.99) | 0.9509 | 29.84 (-6.65,66.33) | 0.1090 |
| Model 2 |  |  | 13.02 (-18.2,44.21) | 0.4130 | -0.79 (-21.1,19.52) | 0.9390 | 27.56 (-7.60,62.73) | 0.1244 |
| Model 3 |  |  | 12.38 (-19.0,43.73) | 0.4388 | -1.43 (-21.8,18.91) | 0.8901 | 28.87 (-6.12,63.86) | 0.1058 |
| Model 4 |  |  | 18.12 (-16.0,52.25) | 0.2980 | -3.65 (-24.9,17.60) | 0.7363 | 28.66 (-6.63,63.95) | 0.1113 |

Presented data are adjusted CWV estimates (95% CI) per 5 µg/m<sup>3</sup> increase for PM<sub>2.5</sub> from a linear mixed model with centre modelled as a random intercept of regio-stratified analysis. Model 1, unadjusted. Model 2, adjusted for age, sex, and ethnicity. Model 3 further adjusted for the INTERHEART risk score and education. Primary model 4 further adjusted for walkability categories.

**Table S5:**
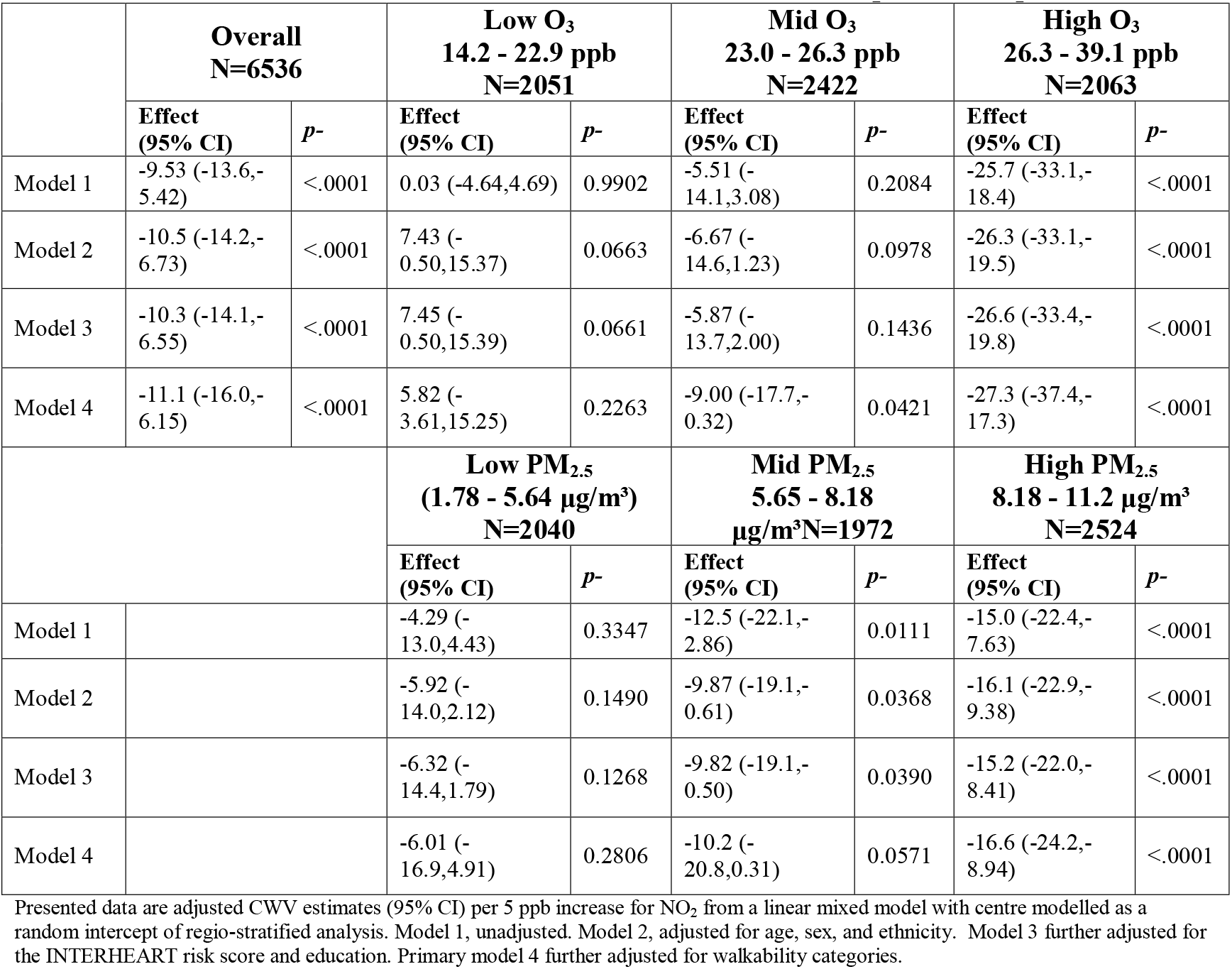
Association of carotid wall volume (mm^3^) with NO_2_ within co-pollutant exposure tertiles.

**Table S6:** Association of carotid wall volume (mm^3^) with pollutant exposure adjusting for neighbourhood socioeconomic status.

| Pollutant | Q1 | Q2 | Q3 | Q4 | Q5 | p-trend |
| --- | --- | --- | --- | --- | --- | --- |
| **PM<sub>2.5</sub>** |  |  |  |  |  |  |
| Quintile range | 1.8 – 5.0 ug/m<sup>3</sup> | 5.0 – 6.0 ug/m<sup>3</sup> | 6.0 – 7.8 ug/m<sup>3</sup> | 7.8 – 8.9 ug/m<sup>3</sup> | 8.9 – 11.2 ug/m<sup>3</sup> |  |
| Model 1 | 911.0 (890.7, 931.3) | 911.4 (891.0, 931.9) | 903.7 (883.6, 923.8) | 902.3 (882.0, 922.6) | 895.3 (874.7, 916.0) | 0.04 |
| Model 2 | 912.8 (889.6, 936.0) | 910.0 (886.8, 933.3) | 902.2 (879.2, 925.2) | 897.8 (874.6, 921.1) | 890.4 (866.9, 913.8) | 0.001 |
| Model 3 | 912.8 (889.9, 935.7) | 909.9 (886.8, 932.9) | 902.0 (879.3, 924.8) | 897.8 (874.9, 920.8) | 890.1 (866.9, 913.3) | 0.001 |
| Model 4 | 909.2 (886.1, 932.3) | 907.3 (884.3, 930.3) | 900.4 (877.7, 923.1) | 898.3 (875.4, 921.2) | 892.3 (868.9, 915.7) | 0.03 |
| Model 5:<br>Model4+ NO<sub>2</sub> | 906.3 (882.7, 929.8) | 904.5 (881.1, 928.0) | 897.3 (874.1, 920.4) | 902.0 (878.6, 925.3) | 896.7 (872.8, 920.5) | 0.29 |
| Model 5:<br>Model4 + O<sub>3</sub> | 908.8 (888.7, 929.0) | 909.0 (888.9, 929.0) | 904.4 (884.6, 924.2) | 905.0 (884.8, 925.3) | 898.7 (878.0, 919.5) | 0.24 |
| **NO<sub>2</sub>** |  |  |  |  |  |  |
| Quintile range | 0.9 – 5.5 ppb | 5.5 – 9.3 ppb | 9.4 – 14.0 ppb | 14.1 – 17.9 ppb | 17.9 – 33.9 ppb |  |
| Model 1 | 920.8 (898.8, 942.8) | 926.8 (906.7, 947.0) | 897.3 (877.9, 916.7) | 900.1 (880.8, 919.5) | 890.9 (871.3, 910.4) | <.0001 |
| Model 2 | 919.9 (896.3, 943.5) | 928.8 (906.7, 950.9) | 895.1 (873.5, 916.6) | 893.9 (872.4, 915.5) | 890.2 (868.5, 911.9) | <.0001 |
| Model 3 | 920.1 (896.6, 943.6) | 928.5 (906.5, 950.5) | 894.8 (873.4, 916.3) | 893.9 (872.5, 915.3) | 890.5 (869.0, 912.1) | <.0001 |
| Model 4 | 919.5 (894.2, 944.8) | 930.3 (907.9, 952.8) | 896.2 (874.6, 917.8) | 892.8 (871.2, 914.4) | 888.7 (866.7, 910.7) | <.0001 |
| Model 5:<br>Model4+O<sub>3</sub> | 916.7 (892.5, 940.9) | 929.6 (908.6, 950.6) | 897.9 (877.6, 918.1) | 896.9 (876.1, 917.6) | 893.2 (872.0, 914.4) | 0.003 |
| Model 5:<br>Model4+PM<sub>2.5</sub> | 918.4 (892.5, 944.3) | 929.8 (906.9, 952.7) | 895.9 (873.9, 918.0) | 892.9 (870.8, 914.9) | 888.9 (866.5, 911.3) | <0.001 |
| **O<sub>3</sub>** |  |  |  |  |  |  |
| Quintile range | 14.2 – 22.0 ppb | 22.1 – 24.4 ppb | 24.4 – 25.7 ppb | 25.7 – 28.0 ppb | 28.0 – 37.1 ppb |  |
| Model 1 | 902.3 (884.4, 920.1) | 898.3 (880.2, 916.4) | 906.6 (888.7, 924.4) | 899.8 (881.5, 918.0) | 925.9 (907.2, 944.6) | 0.02 |
| Model 2 | 898.6 (879.7, 917.5) | 896.8 (877.7, 915.9) | 902.6 (883.7, 921.4) | 897.2 (878.0, 916.4) | 929.6 (910.1, 949.2) | 0.001 |
| Model 3 | 898.4 (879.7, 917.1) | 896.7 (877.9, 915.5) | 903.0 (884.4, 921.6) | 897.4 (878.4, 916.4) | 929.5 (910.1, 948.9) | 0.001 |
| Model 4 | 899.9 (880.3, 919.5) | 897.1 (877.6, 916.7) | 901.6 (882.0, 921.1) | 895.2 (875.2, 915.1) | 924.5 (903.3, 945.6) | 0.03 |
| Model 5:<br>Model4+NO<sub>2</sub> | 903.3 (881.6, 925.0) | 898.4 (876.9, 920.0) | 901.8 (880.2, 923.3) | 891.4 (869.4, 913.4) | 915.3 (891.7, 938.9) | 0.48 |
| Model 5:<br>Model4+PM<sub>2.5</sub> | 899.9 (879.7, 920.1) | 897.1 (876.9, 917.2) | 901.2 (881.0, 921.4) | 894.7 (874.1, 915.3) | 923.3 (901.4, 945.3) | 0.05 |

Presented data are adjusted least square means (95% CI) from a linear mixed model with centre modelled as a random intercept. Model 1, unadjusted. Model 2, adjusted for age, sex, ethnicity. Model 3 further adjusted IHRS & education. Model 4 further adjusted for community level: walkability categories, material factor score & social factor score. Model 5 further adjusted for co-pollutants. P-trend from linear contrasts. For all models, N=6,340.

**Figure S1:**
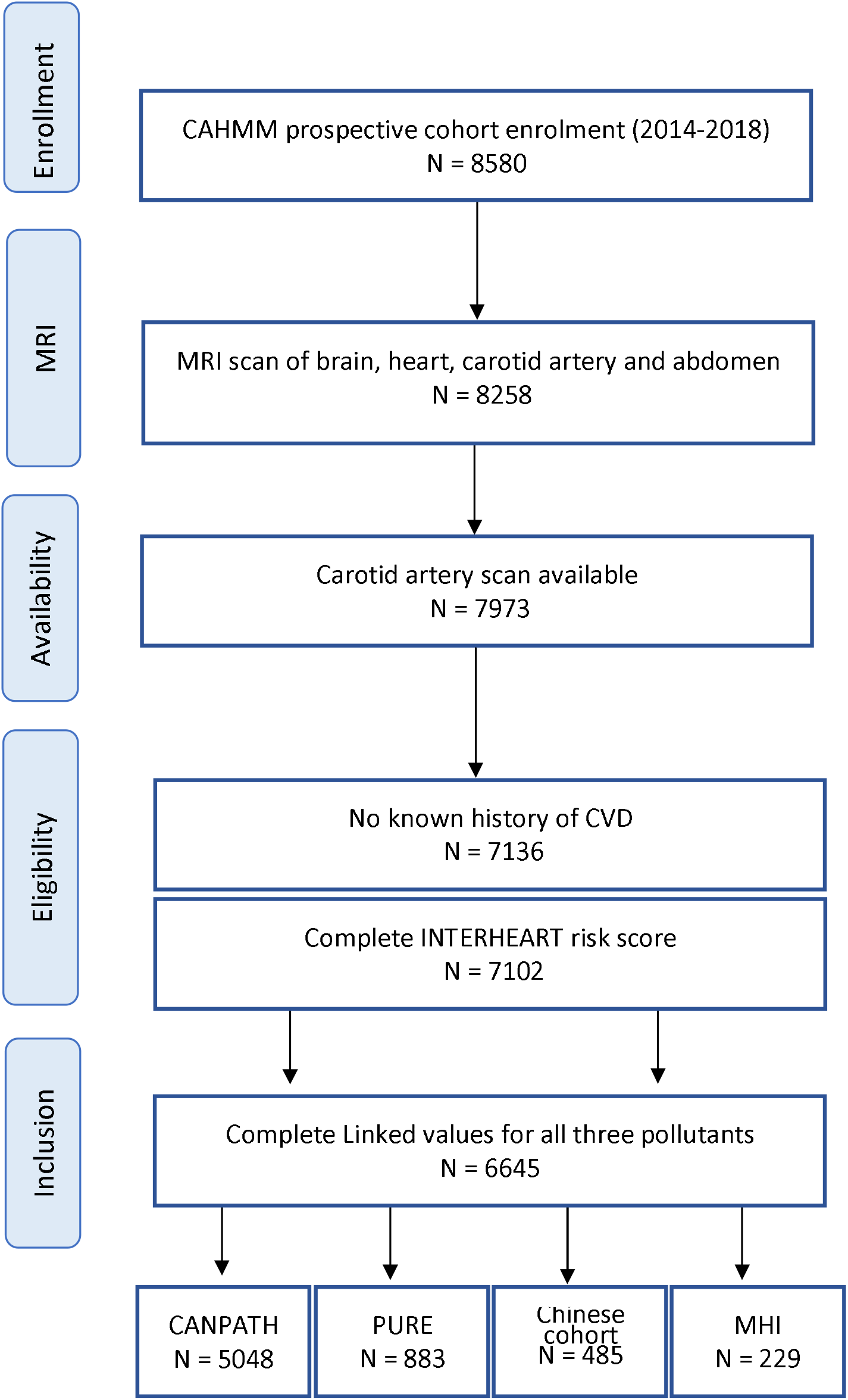
Flow chart for Air pollution and MRI markers in CAHHM.

